# Ethnicity and outcomes in COVID-19 in the United Kingdom: a systematic review and meta-analysis

**DOI:** 10.1101/2021.11.07.21266027

**Authors:** Sania Siddiq, Saima Ahmed, Irfan Akram

## Abstract

This systematic review and meta-analysis evaluated the clinical outcomes of COVID-19 disease in the ethnic minorities of the UK in comparison to the White ethnic group. Medline, Embase, Cochrane, MedRxiv, and Prospero were searched for articles published between May 2020 to April 2021. PROSPERO ID: CRD42021248117. Fourteen studies (767177 participants) were included in the review. In the adjusted analysis, the pooled Odds Ratio (OR) for the mortality outcome was higher for the Black (1.83, 95% CI: 1.21-2.76), Asian (1.16, 95% CI: 0.85-1.57), and Mixed and Other (MO) groups (1.12, 95% CI: 1.04-1.20) compared to the White group. The adjusted and unadjusted ORs of intensive care admission were more than double for all ethnicities (OR Black 2.32, 95% CI: 1.73-3.11, Asian 2.34, 95% CI: 1.89-2.90, MO group 2.26, 95% CI: 1.64-3.11). In the adjusted analysis of mechanical ventilation need the ORs were similarly significantly raised (Black group 2.03, 95% CI: 1.80-2.29, Asian group 1.84, 95% CI: 1.20-2.80, MO 2.09, 95% CI: 1.35-3.22). This review confirmed that all ethnic groups in the UK suffered from increased disease severity and mortality with regards to COVID-19. This has urgent public health and policy implications to reduce the health disparities.

## 1 Introduction

In the UK, there have been 152490 deaths (192 deaths/100000 population), 471045 hospital admissions, and 4717811 confirmed cases (7062 cases per 100000 population) due to COVID-19 pandemic between March 2020 to June 2021 [1]. There have been growing concerns in the UK that the people belonging to the Black, Asian, and Minority Ethnic (BAME) community have been disproportionately impacted by COVID-19 [2-4].

A systematic review (SR) analyzed data between December 2019 to 31st August 2020 and found that Asian people had a higher risk of intensive care admissions and death [5]. However, another review of COVID-19 patients did not find that ethnicity was associated with the worst outcomes [6].

In order to reduce the impact of COVID-19 on the population, COVID-19 related research had been prioritized and classified as urgent public health research by the National Institute for Health Research (NIHR) [7]. Following the government’s call and support for research in this area, numerous studies were conducted in the UK. However, there have not been any published SR and meta-analysis on the impact of COVID-19 on the BAME population in the UK, which has aggregated and synthesized the results of all the newly undertaken studies. Therefore, an up-to-date SR of UK-based studies will quantify the health inequalities faced by the BAME people concerning COVID-19 in the UK. This is necessary to inform policy, promote risk assessment in workplaces and support culturally sensitive public health measures; and prevent the excess avoidable disease burden [6]. The aim of this research is to assess the clinical outcomes of COVID-19 amongst the ethnic minorities in the UK.

## 2 Methods

The review adhered to the Preferred Reporting Items for Systematic Reviews and Meta-Analyses (PRISMA) criteria [8]. The protocol was registered with PROSPERO international prospective register of systematic reviews with the ID. CRD42021248117 [9].

### 2.1 Information Sources

This review was adapted from the systematic review by Sze, Pan [5] and Raharja, Tamara [6]. The reference list of these two key reviews were searched for relevant studies published during the period January 2020 to August 2020. The database searches (Ovid Medline, Ovid Embase, Ovid Cochrane, MedRxiv, and Prospero) were restricted to a one-year period, defined as May 2020 to April 2021.

### 2.2 Search strategy

The Population, Exposure, Comparator, and Outcomes (PECO) framework was used to formulate the criteria for study selection. The population included the adult population aged 18 years and above, in the UK with a confirmed positive COVID-19 result using Reverse Transcriptase-Polymerase Chain Reaction (RT-PCR) tests. The ethnic categorization into White, Black, Asian, Mixed and other groups was based on the UK Census categories 2001 [10]. The Black category included African, Caribbean, Black British, and any other Black background. The Asian category included Indian, Pakistani, Bangladeshi, and any other Asian background. The mixed category included White and Black, White and Asian, or any other mixed background. The other category included Chinese, Arab, and any other backgrounds. The Census 2001 categorization was used because the National Health Service (NHS) utilizes this categorization, which allocates Chinese in the Other group rather than in the Asian group. All-cause mortality and Intensive Care Unit (ICU) admission rates were assessed as the primary outcomes, with Invasive Mechanical Ventilation (IMV) as a secondary outcome. The measures of effect for the outcomes were Hazard Ratio (HR), Risk Ratio (RR), Odds Ratio (OR), or Standardised Mortality Ratio (SMR). Interventional studies, systematic reviews, observational studies including case-control studies, and cohort studies were included. Non-peer-reviewed studies were also included, as this was a rapidly evolving field. Conference abstracts, commentaries, cross-sectional studies, reports, editorials, non-systematic review articles; case reports, late-breaking abstracts, studies without a comparator group, and papers whose full text was unavailable, were excluded. Risk of infection only studies were excluded. Studies were restricted to those in the English language, conducted in the UK, restricted to the adult BAME community, with confirmed COVID-19. Studies from the same population, with similar outcomes, were excluded, as this may have created a duplication of data. Studies that grouped all ethnic minorities as one were excluded, as this grouping would not lead to a meaningful analysis of the burden of illness in the various sub-groups. Specialist librarians were asked to review the search strategy with the keywords of COVID-19, ethnic minority, and the UK. The search strategy was based on the search originally conducted by Sze, Pan [5], and Public Health England (PHE) and was adapted for this review by the addition of ‘UK’ as a key term during the searches [2]. The detailed search strategy for each database is provided in the research proposal [9]. The search terms were tailored for each database and the searches were run separately for each database to enhance sensitivity. The search period defined was between May 2020 to April 2021 and results were limited to English.

### 2.3 Selection process

Two reviewers independently screened the titles and the abstracts of the studies; and excluded non-relevant studies. Full texts of the remaining studies were retrieved and reviewed for inclusion in the study against the selection criteria defined earlier. Any disagreements between the researchers were resolved by discussion.

### 2.4 Data Collection process

One researcher (SA) extracted data from the eligible studies and assessed the risk of bias (ROB). Data extraction was checked by a second reviewer (IA). Authors were also contacted for missing data.

### 2.5 Quality assessment

The Newcastle-Ottawa Scale (NOS) (Supplementary material S1) was employed to assess the ROB in the included studies. A NOS score of 7-9 is classed as low ROB, a score of less than 5 as high ROB; and a score of 5-6 as moderate ROB [11]. One reviewer (SA) carried out the quality assessment which was at the study level and the outcome level using the Grading of Recommendations Assessment, Development and Evaluation (GRADE) approach [12]. Publication bias was not assessed as there were too few studies in the adjusted analysis.

### 2.6 Data synthesis

The raw counts for the various outcome variables were used to calculate the RR and 95% Confidence Intervals (CIs). A meta-analysis was conducted for the outcomes which compared risks in Black, Asian, or Mixed and Others (MO) groups; with the White group in at least two studies. Studies that did not use the White group as a comparator group were excluded from the meta-analysis. As all the studies were observational designs, a Der Simonian-Laird Random-Effects Meta-analysis (REM) was conducted for all outcomes as per the Cochrane recommendations due to the heterogeneous nature of these studies [13]. For rare outcomes, OR was assumed to be equal to RR and RR was assumed to be equal to HR [5, 6]. The studies which provided Adjusted Odds Ratio (AOR) were pooled together in one group and those which provided Adjusted Hazard Ratio (AHR) were separately pooled together in another group (if the outcomes were not rare). In studies where the raw data was missing, the authors were contacted to obtain this. In order to include studies with missing data in the meta-analysis, the unadjusted HR/RR were combined using the inverse variance method. Using this method, the pooled risk estimates were calculated separately for each ethnic group and a summary statistic was provided. The results were written in tabulated form and as forest plots. Excel and RevMan were used to analyze and tabulate the data. Origin 2021b was used to convert graphical data to tabulated form as advised by Cochrane to avoid mistakes in the manual conversion of data [14]. The statistical heterogeneity was explored by calculating the I^2^ statistic using RevMan and by looking at the overlap of the CIs in the forest plots. A subgroup analysis was conducted to explore differences in risk estimates across the subgroups [15]. Studies based on ICU patients only, general hospital patients only, and the general population were analyzed separately. A sensitivity analysis was conducted based on only peer-reviewed studies and a separate analysis was conducted based on studies with low ROB [15].

## 3 Results

The search on Medline, Embase, MedRxiv, Cochrane, and Prospero yielded 939 studies on 14-15 April 2021. In the reference lists of the two key SRs, 147 references were found [5, 6]. After the removal of duplicates, 849 results were left. Seven hundred and ninety-four results were excluded, and 63 studies were selected for full-text review. Forty-nine studies were excluded with reasons as shown in Figure 1 and Supplementary Table S1. Fourteen studies were selected to be included in the systematic review.

**Figure 1:**
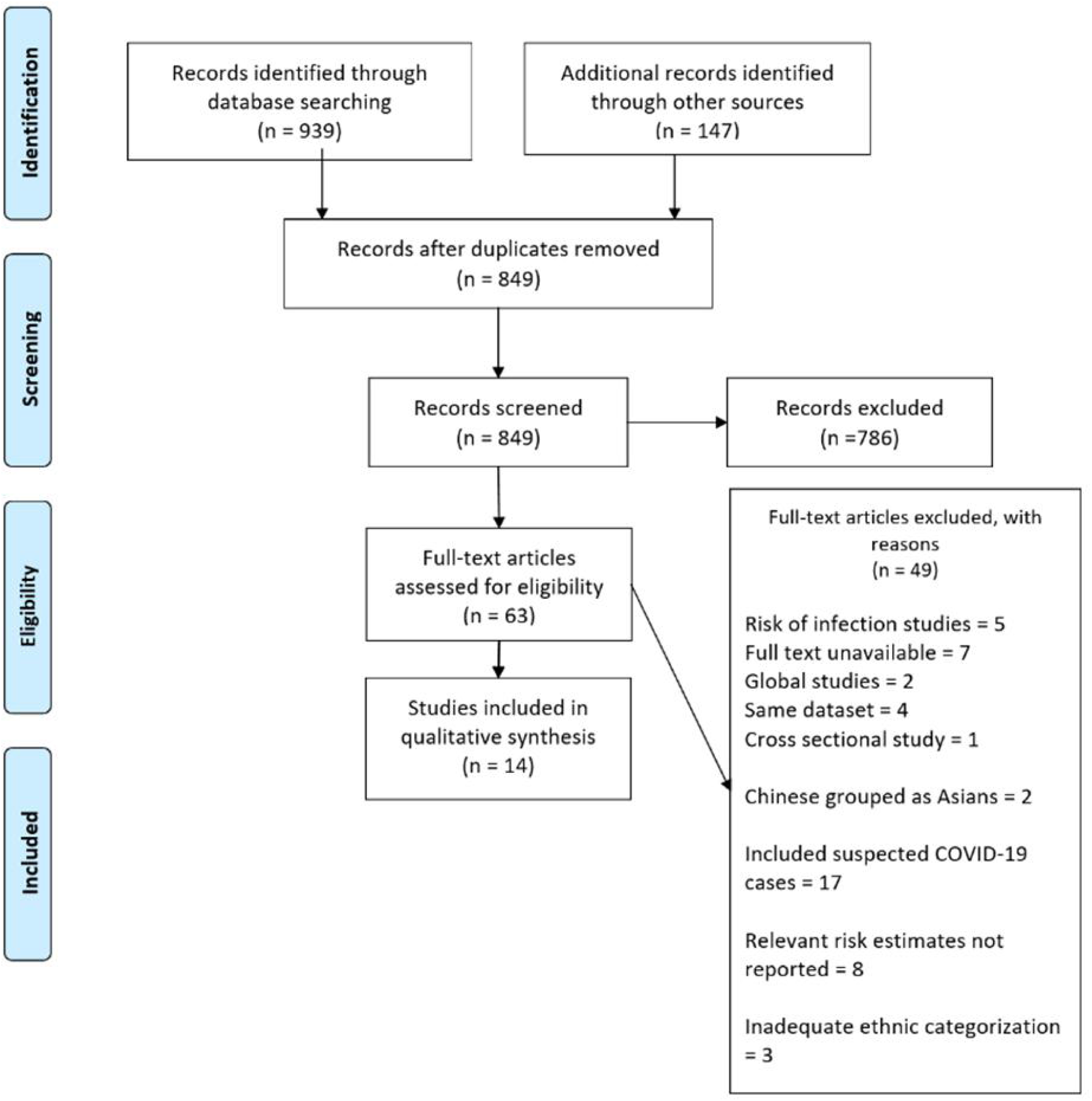
PRISMA flow diagram [8]

### 3.1 Study characteristics

The 14 studies included a total of 767177 participants. All the studies were based in the UK. The study design, population type, setting, sample size, outcomes, comorbidities, and confounders adjusted for are given in Table 1 [16-29]. The sample size in this review varied from 140 to 448664.

**Table 1:**
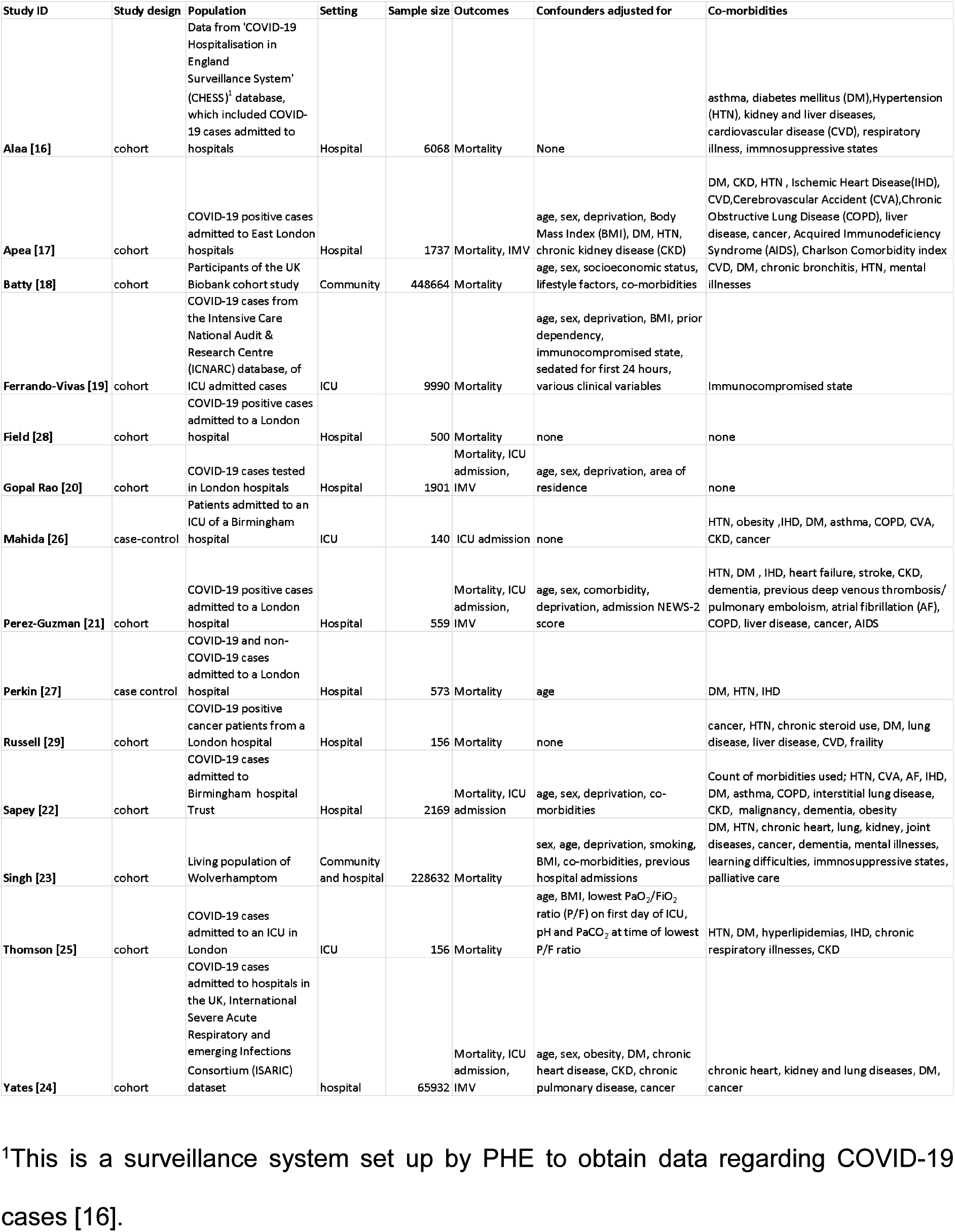
Study characteristics of included studies

Two studies were based on community participants and the rest were based on hospital settings. Two studies were not peer-reviewed [18, 28]. Fourteen studies were included in the systematic review and 12 in the meta-analysis. Two studies did not use White as a comparator group and so these were not included in the meta-analysis [26, 27]. Twelve studies were cohort studies, and two were case-control studies. Thirteen studies provided data on risks of mortality due to COVID-19 in the various ethnic groups, six on ICU admission, and four on IMV need.

### 3.2 Quality of studies

The risk of bias (ROB) was low, average NOS score was 7 (ranged from 4-9). Nine studies had a low ROB score (green color) on the NOS, four studies had a moderate ROB (yellow color) score on the NOS and one study had a high ROB (red color) as shown in Table 2. The low scores were largely due to studies that failed to adjust for confounding factors. Perez-Guzman, Daunt [21] provided raw outcome data about the number of cases admitted to the ICU and who received IMV, but did not perform any statistical analysis of this data. Similarly, Apea, Wan [17] provided raw data about ICU admissions, but no risk estimates were provided. One study had a high ROB and did not use White as a comparator group and so was excluded from the meta-analysis [26].

**Table 2:** Quality assessment of studies using a modified NOS for assessing studies

| Study ID | Selection | Selection | Selection | Selection | Comparability | Outcome |  | Outcome | Total (*9) |
| --- | --- | --- | --- | --- | --- | --- | --- | --- | --- |
|  | Representativeness<br>of exposed cohort (*) | Selection<br>of non-<br>exposed<br>cohort (*) | Ascertainment<br>of exposure (*) | Outcome<br>not present<br>at start (*) | (**) | Assessment<br>of outcome<br>(*) | Follow-<br>up long<br>enough<br>(*) | Adequacy<br>of follow-<br>up (*) |  |
| Alaa [16] | * | * | * | * |  | * | * |  | 6 |
| Apea [17] | * | * |  | * | ** | * | * | * | 8 |
| Batty [18] | * | * |  | * | ** | * | * |  | 7 |
| Ferrando-Vivas [19] | * | * | * | * | * | * | * | * | 8 |
| Field [28] | * | * | * | * |  | * | * |  | 6 |
| Gopal Rao [20] | * | * | * | * | * | * | * | * | 7 |
| Mahida [26] |  | * | * | * |  | * |  |  | 4 |
| Perez-Guzman [21] | * | * | * | * | ** | * | * | * | 9 |
| Perkin [27] | * | * |  | * | * | * | * |  | 6 |
| Russell [29] |  | * | * | * |  | * | * |  | 5 |
| Sapey [22] | * | * | * | * | ** | * | * | * | 9 |
| Singh [23] | * | * | * | * | ** | * | * | * | 9 |
| Thomson [25] |  | * | * | * | * | * | * | * | 7 |
| Yates [24] | * | * | * | * | ** | * | * | * | 9 |

Colour coding: green for high, yellow for medium, and red for low NOS

### 3.3 Mortality

The unadjusted OR, RR, AOR, and AHR are shown in Figures 2-5. In the unadjusted analysis, the risk of death was similar in Blacks and Asians (Black OR: 0.89, 95% CI: 0.71-1.12, I^2^=83%, number of studies k=9) (Asian OR: 0.83, 95% CI: 0.68-1.02, I^2^=85%, k=9), but significantly reduced in Mixed and Others group (OR: 0.64, 95% CI: 0.55-0.74, I^2^= 42%, k=9). The adjusted mortality risk was significantly raised for the Asian group (1.32, 95% CI: 1.22-1.42, I^2^= 0%, k=3) but not for the Black and Mixed and Others groups. The odds of dying were significantly increased for the Blacks, Mixed and Other ethnicities compared to the White group in the adjusted analysis (Black AOR: 1.83, 95% CI: 1.21-2.76, I^2^= 87%, k=6, MO AOR: 1.12, 95% CI: 1.04-1.20, I^2^= 0%, k=5) groups, but not for the Asian ethnic group. All the studies in the adjusted analysis were low ROB studies. In the sensitivity analysis, with only published studies, the increased odds of mortality in the Black and Mixed and Others groups was maintained (Black AOR 1.48, 95% CI: 1.10-1.99, and MO AOR 1.12, 95% CI: 1.04-1.20) as shown in Table 3. The odds of increased mortality for all ethnicities was stronger in subgroup analysis with only hospital-based studies (AOR= 1.22, 95%CI 1.07-1.38, I^2^=6%, k=4 for Blacks, AOR= 1.28, 95%CI 1.04-1.57, I^2^=40%, k=4 for Asians, AOR= 1.12, 95%CI 1.04-1.20, I^2^=0%, k=4 for MO). This subgroup had low heterogeneity and showed statistically significant results as shown in Table 4. In the adjusted analysis, of only population-based studies, the odds of dying for the Black ethnic group was almost three times that of the White ethnic group (AOR=2.94, 95% CI: 1.46-5.90), but with a larger CI. This was similar to the Office for National Statistics (ONS) data [30]. However, the odds were not raised for the Asian group. Perkin, Heap [27] found that adjusted odds of mortality due to COVID-19 were increased for all ethnicities (Asian AOR 3.62, 95% CI=1.84–7.11, Black AOR 2.91, 95% CI= 1.43–5.91, and Other AOR 3.01, 95% CI=1.61–5.64) compared with hospital deaths in 2019.

**Table 3:** Sensitivity analysis for the mortality outcome

|  | Studies | Pooled unadjusted<br>OR (95% CI) | I<sup>2</sup> | Studies | Pooled unadjusted<br>RR (95% CI) | I<sup>2</sup> | Studies | Pooled AOR (95% CI) | I<sup>2</sup> |
| --- | --- | --- | --- | --- | --- | --- | --- | --- | --- |
| **All studies with raw mortality data** |  |  |  |  |  |  |  |  |  |
| Black | 9 | 0.89, (0.71- 1.12) | 83% | 11 | 0.98 (0.86- 1.12) | 88% | 6 | **1.83, (1.21-2.76)** | 87% |
| Asian | 9 | 0.83, (0.68- 1.02) | 85% | 11 | 0.97 (0.84- 1.12) | 91% | 6 | 1.16, (0.85-1.57) | 71% |
| Mixed and Other | 9 | **0.64, (0.55- 0.74)** | 42% | 10 | **0.74 (0.64- 0.85)** | 74% | 5 | **1.12, (1.04-1.200)** | 0% |
| **Only published, peer-reviewed studies** |  |  |  |  |  |  |  |  |  |
| Black | 9 | 0.89 (0.71- 1.12) | 83% | 10 | 0.98 ( 0.83- 1.15) | 84% | 5 | **1.48 ( 1.10- 1.99)** | 69% |
| Asian | 9 | 0.83 (0.68- 1.02) | 85% | 10 | 0.96 ( 0.82- 1.140) | 90% | 5 | 1.14 ( 0.80- 1.60) | 77% |
| Mixed and Other | 9 | **0.64 (0.55-0.74)** | 80% | 10 | **0.74 (0.64 - 0.85)** | 45% | 4 | **1.12 ( 1.04 - 1.20)** | 0% |
| **Only studies with low ROB** |  |  |  |  |  |  |  |  |  |
| Black | 8 | 0.92 (0.73- 1.15) | 84% | 8 | 0.94 (0.80- 1.11) | 85% | 6 | **1.83 ( 1.21- 2.76)** | 87% |
| Asian | 8 | **0.81 (0.66- 0.99)** | 86% | 8 | 0.86 (0.74- 1.00) | 88% | 6 | 1.16 (0.85- 1.57) | 71% |
| Mixed and Other | 8 | **0.63 (0.54- 0.74)** | 48% | 8 | **0.72 (0.64- 0.82)** | 58% | 6 | **1.12 (1.04- 1.20)** | 0% |

**Table 4:** Subset analysis for the mortality outcome

|  | Studies | Pooled unadjusted<br>OR (95%CI) | I<sup>2</sup> | Studies | Pooled unadjusted<br>RR (95%CI) | I<sup>2</sup> | Studies | Pooled AOR (95%CI) | I<sup>2</sup> |
| --- | --- | --- | --- | --- | --- | --- | --- | --- | --- |
| **All studies with raw mortality data** |  |  |  |  |  |  |  |  |  |
| Black | 9 | 0.89 (0.71- 1.12) | 83% | 11 | 0.98 (0.86- 1.12) | 88% | 6 | **1.83, (1.21-2.76)** | 87% |
| Asian | 9 | 0.83 ( 0.68- 1.02) | 85% | 11 | 0.97 (0.84- 1.12) | 91% | 6 | 1.16, (0.85-1.57) | 71% |
| Mixed and Other | 9 | **0.64 ( 0.55- 0.74)** | 42% | 10 | **0.74 (0.64- 0.85)** | 74% | 5 | **1.12, (1.04-1.20)** | 0% |
| **Hospital based** |  |  |  |  |  |  |  |  |  |
| Black | 8 | 0.82 (0.67- 1.00) | 75% | 8 | 0.87 (0.75- 1.01) | 79% | 4 | **1.22 (1.07 to 1.38)** | 6% |
| Asian | 8 | 0.87 (0.70- 1.09) | 86% | 8 | 0.94 (0.80-1.11) | 89% | 4 | **1.28 (1.04 to 1.57)** | 40% |
| Mixed and Others | 8 | **0.68 (0.60- 0.76)** | 23% | 8 | **0.76, (0.69- 0.84)** | 33% | 4 | **1.12 ( 1.04 to 1.20)** | 0% |
| **Population based** |  |  |  |  |  |  |  |  |  |
| Black |  |  |  |  |  |  | 2 | **2.94, (1.46-5.90)** | 83% |
| Asian |  |  |  |  |  |  | 2 | 0.80, (0.30-2.15) | 81% |
| Mixed and Others |  |  |  |  |  |  |  |  |  |
| **Intensive care based** |  |  |  |  |  |  |  |  |  |
| Black | 2 | 1.00, (0.87-1.15) | 0% | 2 | 1.00 (0.92- 1.09) | 0% |  |  |  |
| Asian | 2 | 1.36, (0.65- 2.80) | 67% | 2 | 1.24 (0.74- 2.08) | 69% |  |  |  |
| Mixed and Others | 2 | 0.74 (0.51-1.08) | 5% | 2 | 0.82 (0.57- 1.17) | 6% |  |  |  |

**Figure 2:**
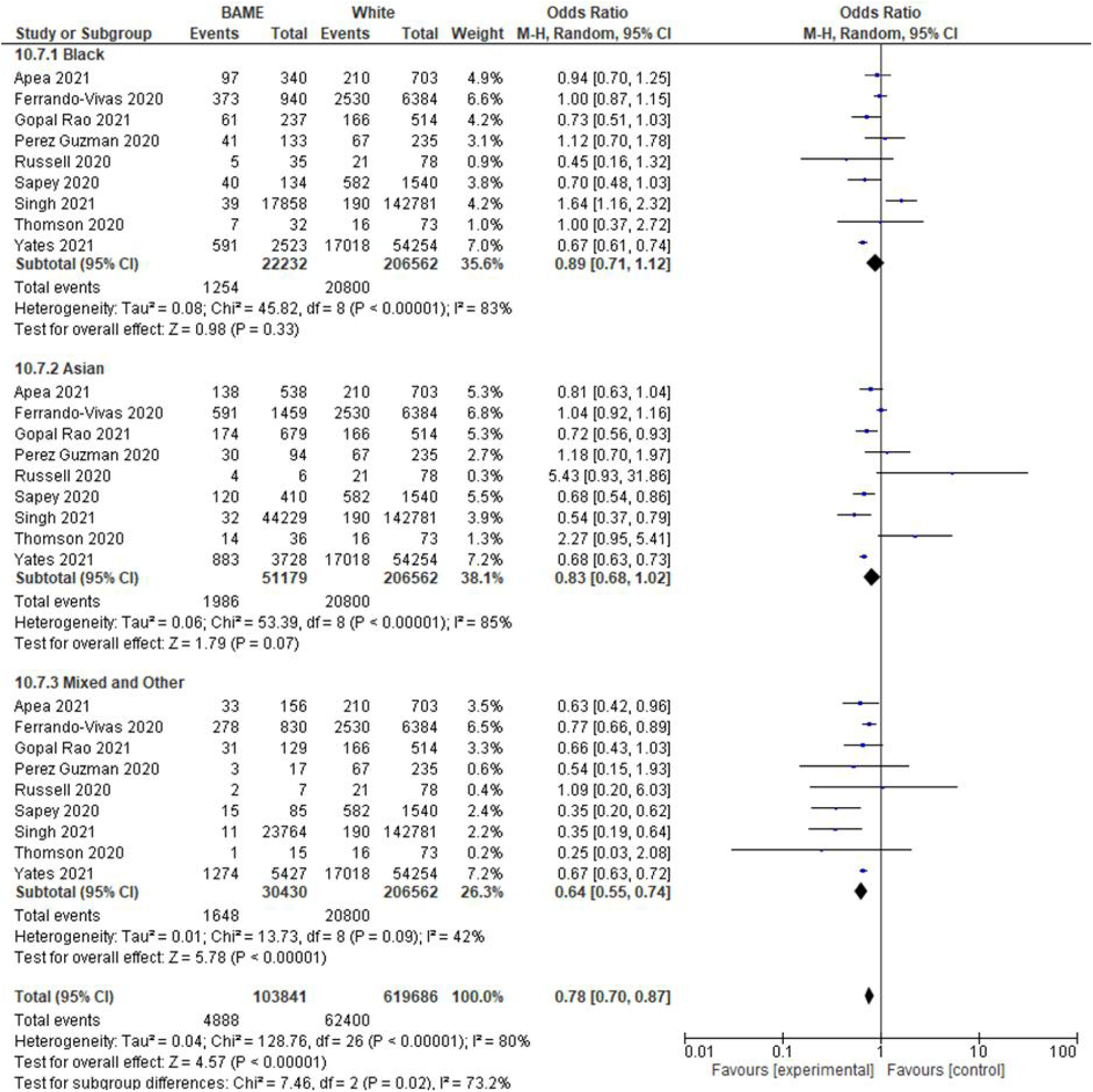
Forest plot of unadjusted OR for the mortality outcome

**Figure 3:**
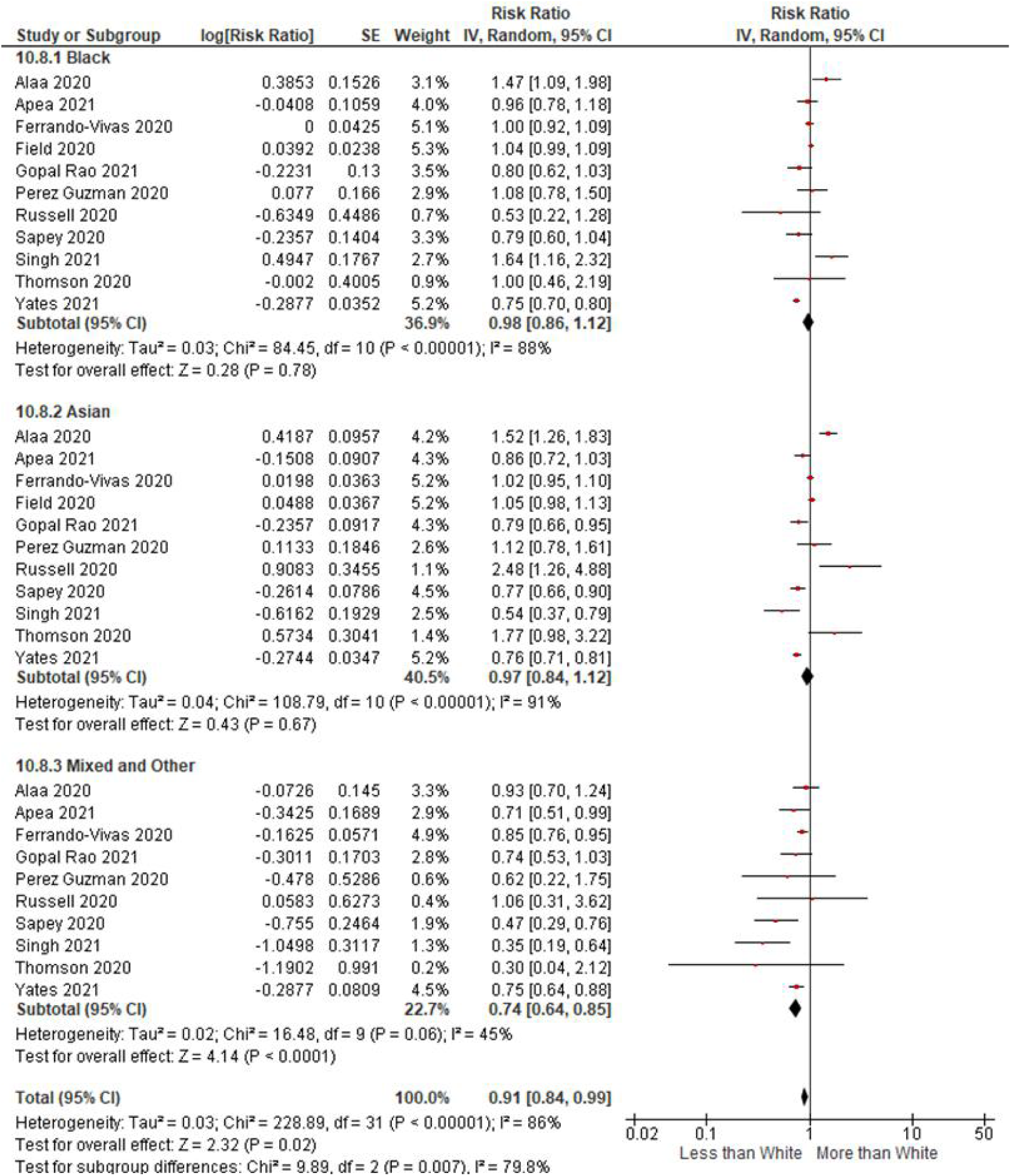
Forest plot of unadjusted RR for the mortality outcome (REM, Inverse variance method)

**Figure 4:**
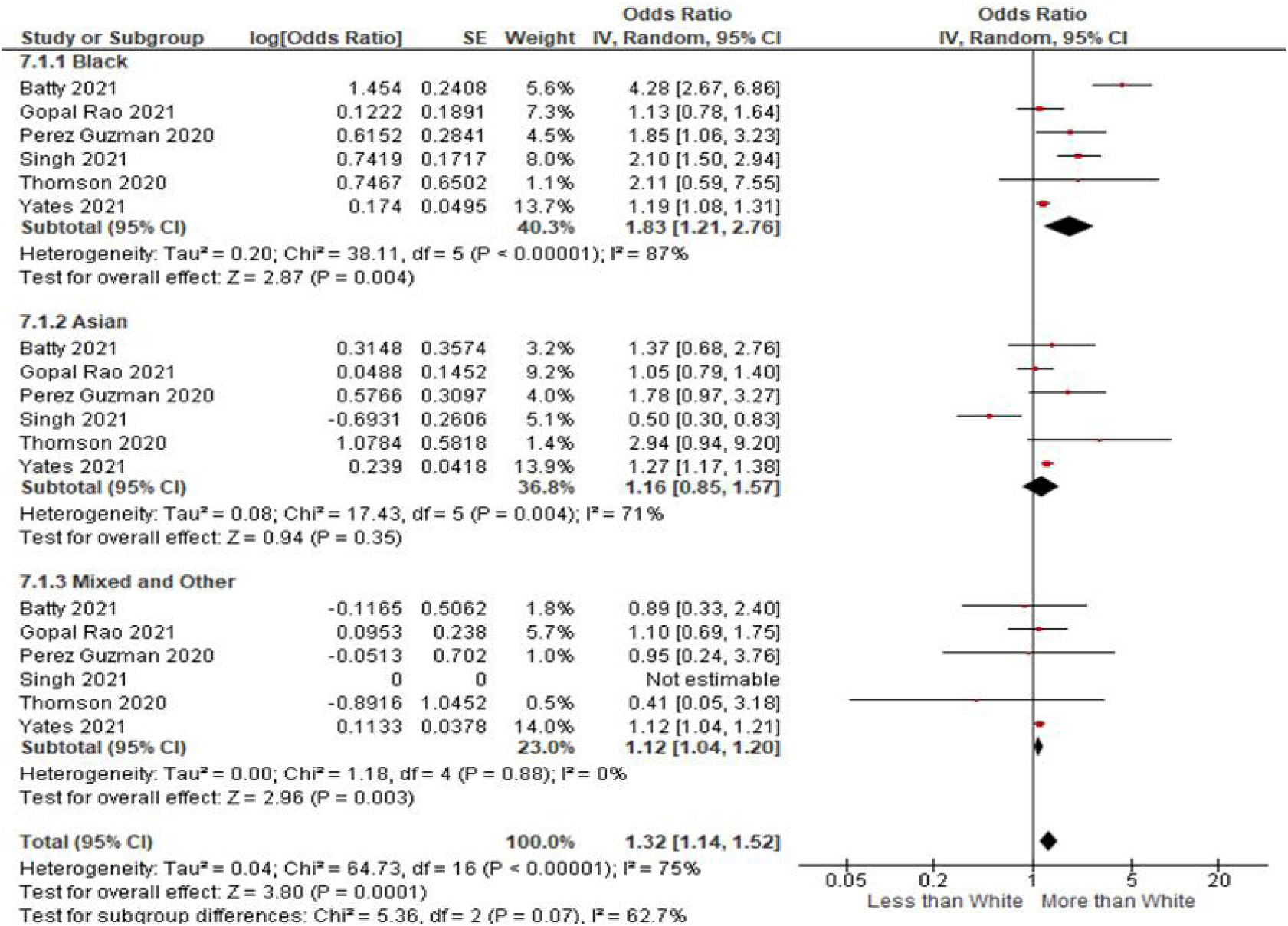
Forest plot of AOR, REM, for the mortality outcome

**Figure 5:**
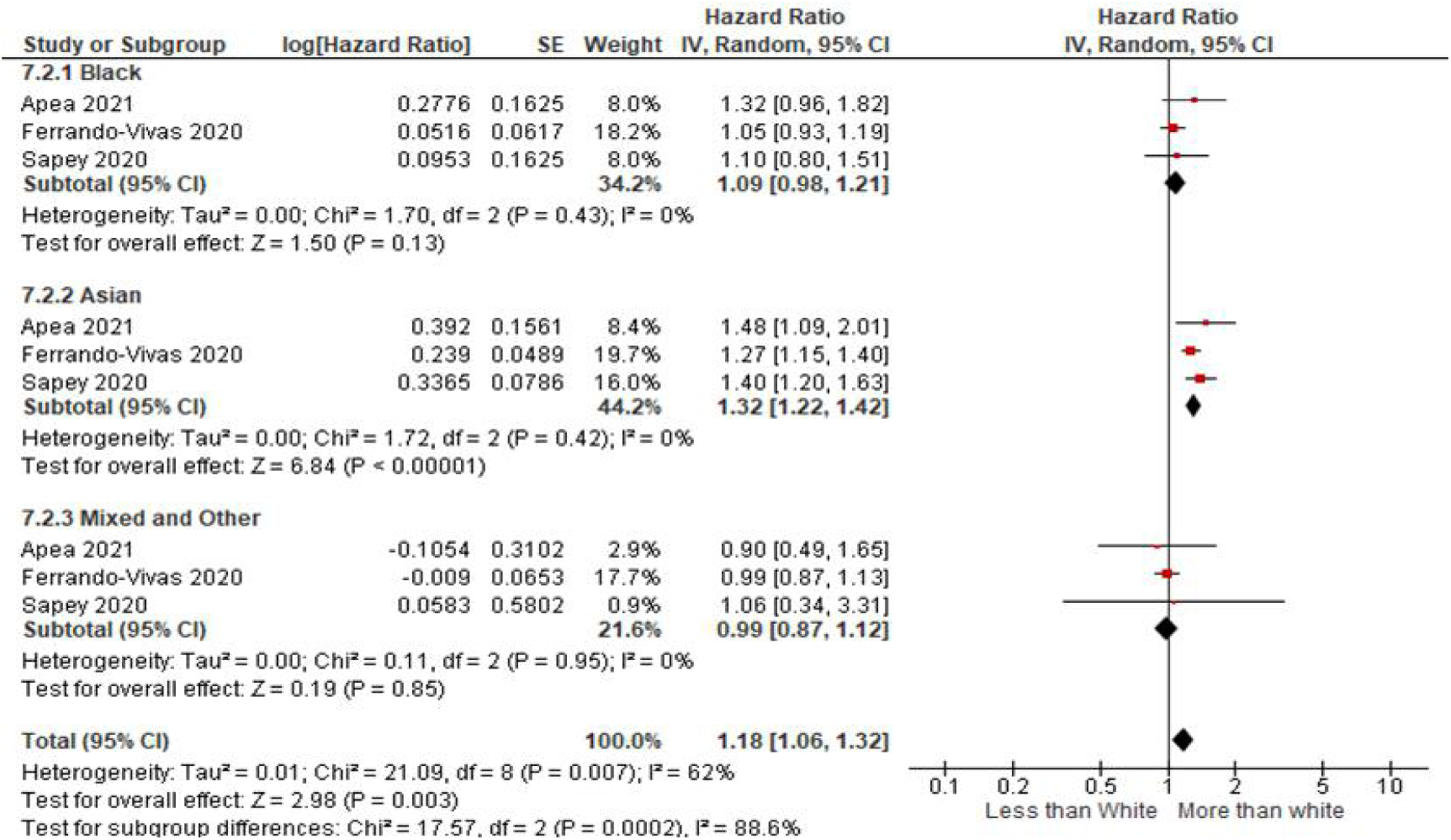
Adjusted HR (Inverse variance method) mortality forest plot

### 3.4 Intensive care admission

Six studies provided data about ICU admissions for the various ethnic groups. Five of these studies were suitable for aggregating the raw outcomes and for pooling the unadjusted risk estimates. These studies included a total of 71791 participants who were admitted to critical care units in the UK. Eighty per cent of them were White, 5% were Black, 9.5% were Asians and 8% were MO. The unadjusted and adjusted analyses for ICU admission are shown in Figures 6 and 7. In the adjusted and unadjusted analysis, the odds of ICU admissions were more than double for patients of all ethnicities as compared to Caucasians. The unadjusted OR for the Black group was 2.32 (95% CI: 1.73-3.11, I^2^= 66%, k=5), for the Asian group it was 2.34 (95% CI: 1.89-2.90, I^2^= 58%, k=5), and for the MO group it was 2.26 (95% CI: 1.64-3.11, I^2^= 45%, k=4). In the pooled AOR, the results were not statistically significant for all ethnicities as the lower CI crossed the line of no effect as shown in Figure 7. However, the results indicated a strong association (OR twice as high) between ethnicity and ICU admission outcome. The pooled AOR for the Black group was 2.61 (95% CI: 0.89-7.68, I^2^= 91%, k=2), for the Asian group it was 2.05 (95% CI: 0.85-4.94, I^2^= 89%, k=2), and for the MO group it was 2.12 (95% CI: 0.94-4.78, I^2^= 80%, k=2). One study which was not included in the meta-analysis compared patients admitted to ICU with COVID-19, and patients admitted to ICU with community-acquired pneumonia (non-COVID controls) [26]. The study found that the cases with COVID-19 had statistically significantly fewer White (p= 0.012) and more Asian cases (p= 0.002) [26].

**Figure 6:**
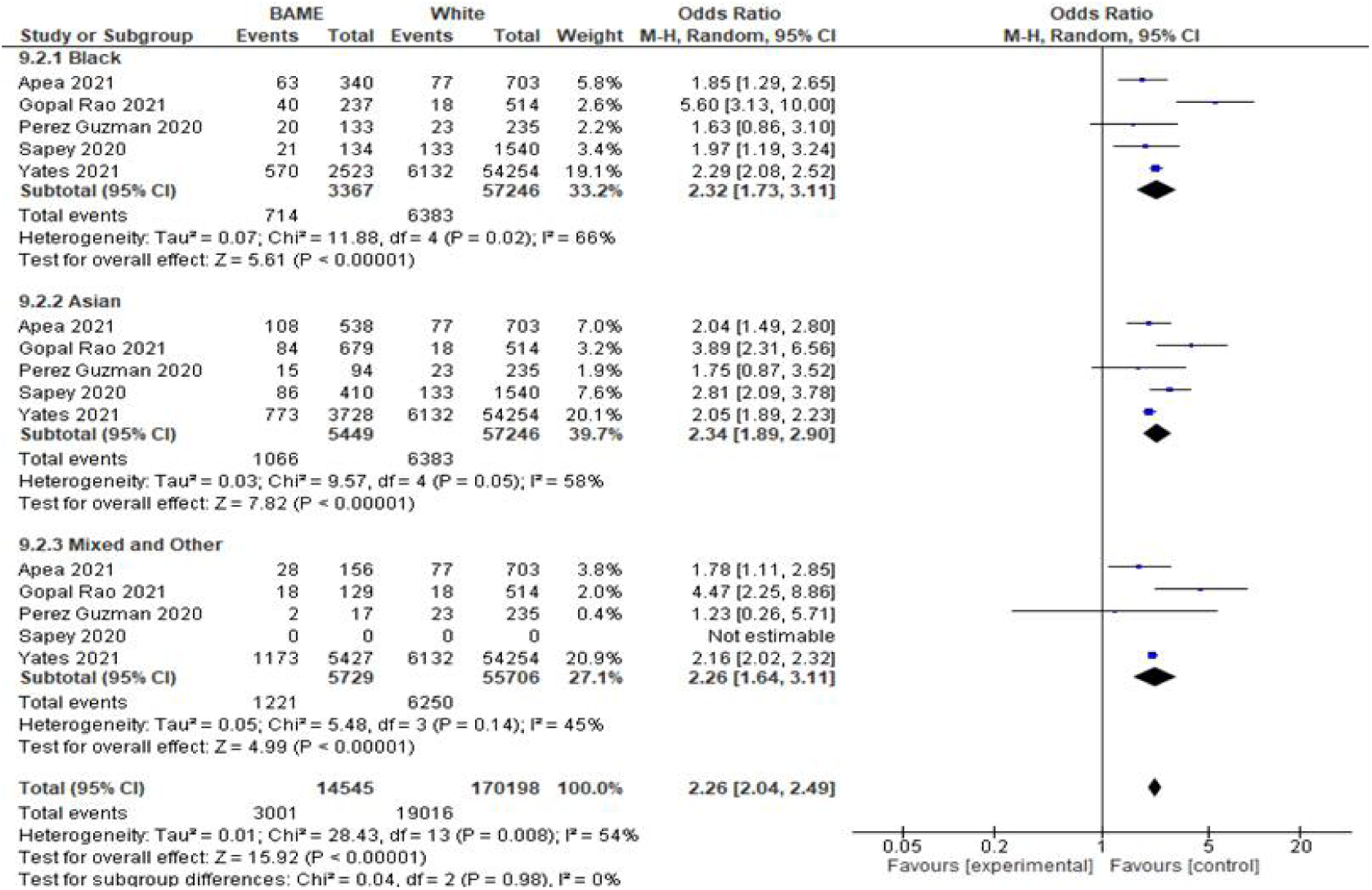
Forest plot of the ICU admission outcome, pooled unadjusted OR, REM

**Figure 7:**
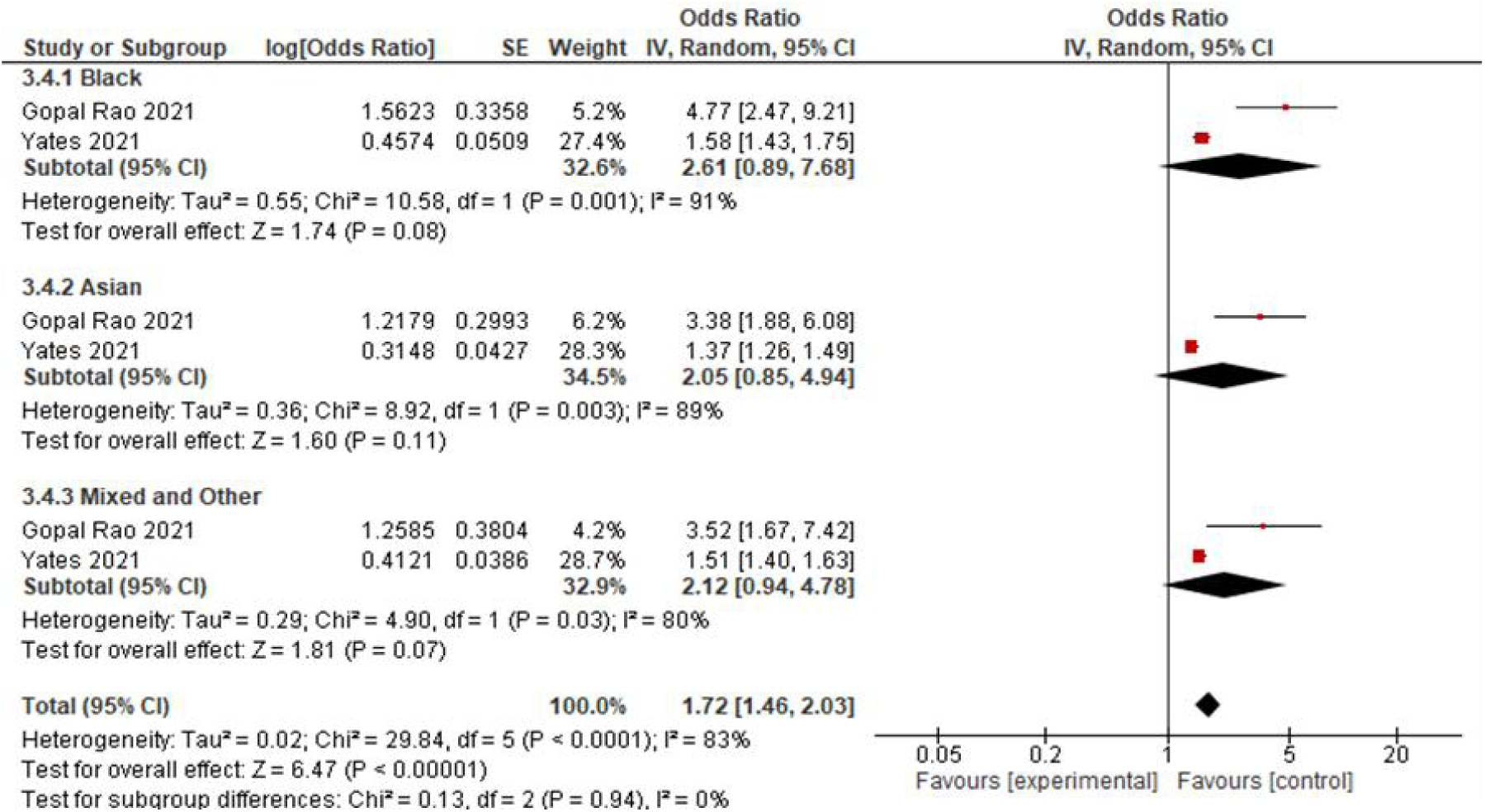
Forest plot of ICU admission outcome, pooled AOR, REM

### 3.5 Mechanical ventilation

Four cohort studies reported ethnicity data about the need for IMV for hospitalized patients in the UK. Amongst a total of 69707 patients, 80% were White, 5% were Black, 7% were Asian and 8% were from MO ethnic group. In the unadjusted analysis, the odds for the Black, Asian, and Mixed and Others were twice more, compared to Whites (Black OR: 2.44, 95% CI: 1.67-3.57, I^2^=67%, k=4, Asian OR: 2.29, 95% CI: 1.69-3.11, I^2^=58%, k=4, and the MO groups OR: 2.67, 95% CI: 1.77-4.01, I^2^=53%, k=4) as shown in Figure 8. After adjusting for confounders, the odds of needing ventilation were still raised for all ethnicities, indicating that other factors may be putting them at increased risk. The AOR for the Black group was 2.03 (95% CI: 1.80-2.29, I^2^= 1%, k=3), for the Asian group it was 1.84 (95% CI: 1.20-2.80, I^2^= 74%, k=3) and for the MO group it was 2.09 (95% CI: 1.35-3.22, I^2^= 60%, k=3) as shown in Figure 9.

**Figure 8:**
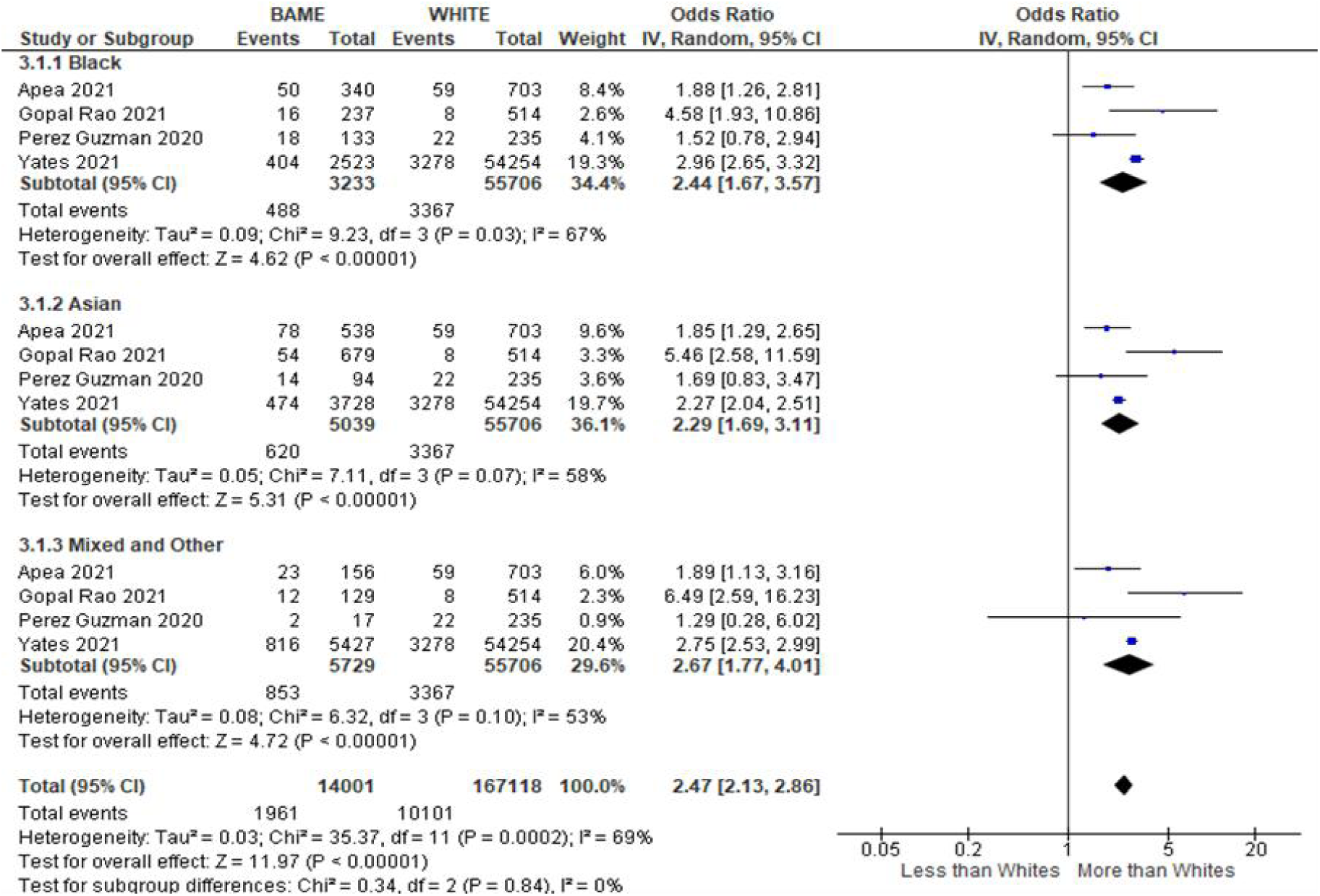
Forest plot of IMV outcome, pooled unadjusted OR, REM

**Figure 9:**
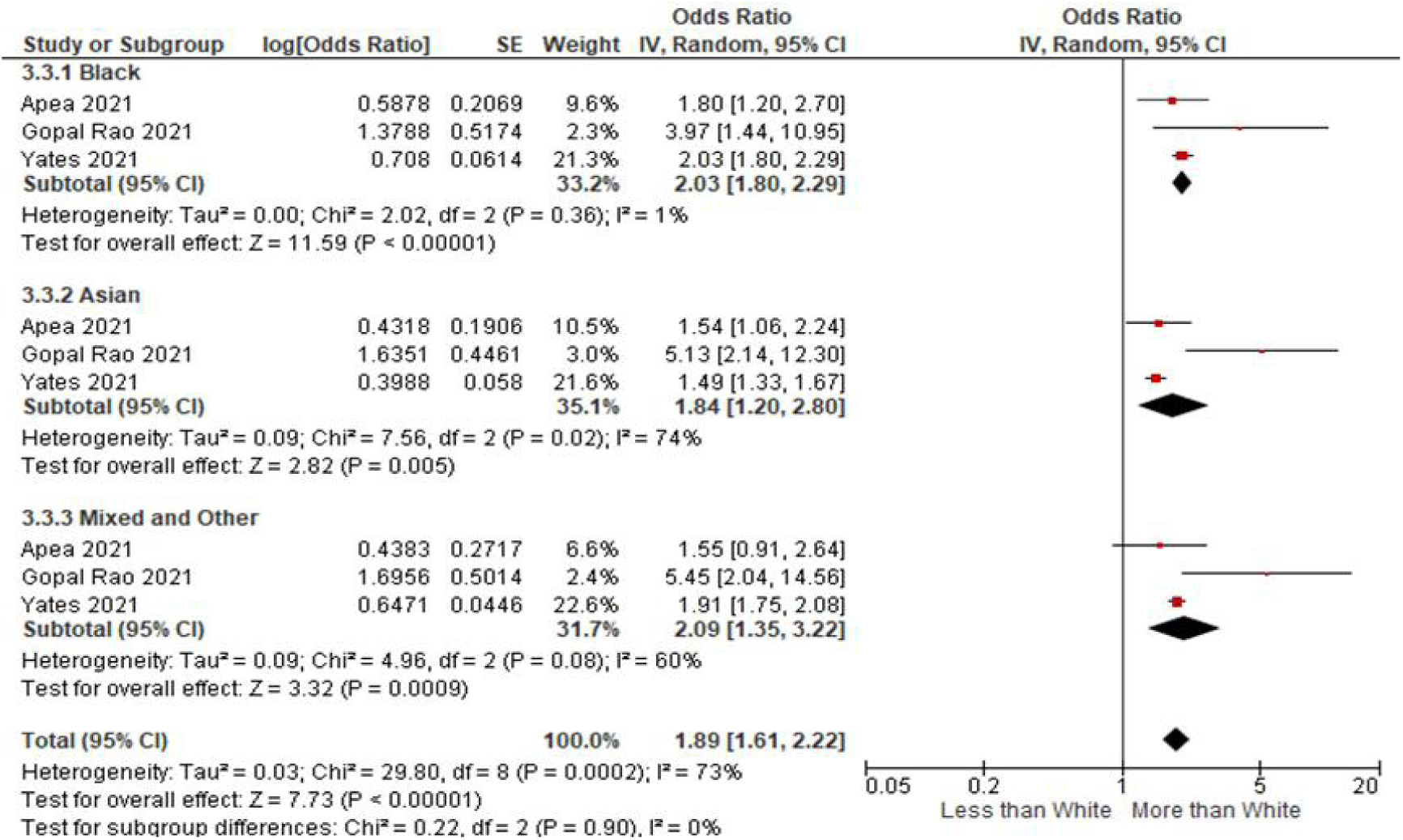
Forest plot of the IMV outcome, pooled adjusted OR, REM

The studies which were included in the IMV need and ICU admission analysis are all low ROB studies, published, and hospital-based studies and so further sensitivity analyses were not conducted. The heterogeneity was high for most of the outcomes.

### 3.6 Quality of evidence assessment

As best evidence regarding risk factors is usually obtained from observational studies, so the evidence in this review was started with high ratings as advised by Foroutan, Guyatt [31] (for prognostic studies). The overall GRADE assessment indicated a high level of confidence for all outcomes, except for the mortality outcomes for the Asian group which was moderate and was downrated due to inconsistency, as shown in the summary of findings in Table 5.

**Table 5:** GRADE, a summary of findings table

| MORTALITY |  |  |  |  |  |  |  |  |  |
| --- | --- | --- | --- | --- | --- | --- | --- | --- | --- |
|  | AOR | Number of participants (studies) | ROB | Inconsistency | Imprecision | Indirectness | Publication bias | Strong association | GRADE |
| BLACK | 1.83 (1.21-2.76)<br>P=0.00001, I<sup>2</sup>=87% | 745844 (6) | no | no | no | no | - | potential risk factor, OR between 1-2 | high |
| ASIAN | 1.16 (0.85-1.57)<br>P=0.004, I<sup>2</sup>=71% | 745844 (6) | no | yes | no | no | - | neutral | moderate, due to inconsistency |
| MO | 1.12 (1.04-1.20)<br>P=0.88, I<sup>2</sup>=0% | 745844 (6) | no | no | no | no | - | potential risk factor, OR between 1-2 | high |
| ICU ADMISSION |  |  |  |  |  |  |  |  |  |
|  | AOR | Number of participants (studies) | ROB | Inconsistency | Imprecision | Indirectness | Publication bias | Strong association | GRADE |
| BLACK | 2.61 (0.89-7.68)<br>P=0.001, I<sup>2</sup>=91% | 67833 (2) | no | no | no | no | - | yes | high, due to strong association |
| ASIAN | 2.05 (0.85-4.94)<br>P=0.003, I<sup>2</sup>=89% | 67833 (2) | no | no | no | no | - | yes | high, due to strong association |
| MO | 2.12 (0.94-4.78)<br>P=0.03, I<sup>2</sup>=80% | 67833 (2) | no | no | no | no | - | yes | high, due to strong association |
| IMV |  |  |  |  |  |  |  |  |  |
|  | AOR | Number of participants (studies) | ROB | Inconsistency | Imprecision | Indirectness | Publication bias | Strong association | GRADE |
| BLACK | 2.03 (1.8-2.29)<br>P=0.36, I<sup>2</sup>=1% | 69570 (3) | no | no | no | no | - | yes | high, due to strong association |
| ASIAN | 1.84 (1.2-2.8)<br>P=0.02, I<sup>2</sup>=74% | 69570 (3) | no | no | no | no | - | no | high, due to strong association |
| MO | 2.09 (1.35-3.22)<br>P=0.08, I<sup>2</sup>=60% | 69570 (3) | no | no | no | no | - | yes | high, due to strong association |

Colour coding: green for high, yellow for medium, and red for low

## 4 Discussion

There was heterogeneity in the populations, settings, methodology, and statistical analysis. The UK is varied in the ethnic distribution of its population. A few studies were conducted in areas of East London, Birmingham, and Wolverhampton, which are very ethnically diverse areas, are comparatively different to the rest of the UK and may have contributed to the clinical heterogeneity in the sample. The meta-analysis was still conducted as this degree of heterogeneity had been reported by other reviews and conducting a meta-analysis was still beneficial.

The adjusted mortality analysis, adjusted population subset analysis, and adjusted hospital subset analysis all indicate raised odds of mortality for all ethnicities. The AOR showed a strong association between the IMV need and ICU admission outcomes for all ethnicities (in Figures 7 and 9). This indicates greater disease severity in all the ethnic minorities necessitating ICU admission and IMV provision. Overall, it can be said that ethnicity is a risk factor for worse prognosis in all the ethnic groups and they do suffer from increased disease severity. Several studies and data from Office for National Statistics (ONS) and Intensive Care National Audit and Research Centre (ICNARC) also validate this finding of worst outcomes of COVID-19 in the ethnic minorities in the UK [2, 5, 30, 32-35].

### 4.1 Strengths

This is the first SR and meta-analysis conducted to assess the burden of disease faced by the BAME community in the UK. The strength of the research was a comprehensive search on relevant databases for published and pre-print articles. A systematic process and a meta-analysis strengthened and clarified the results in the context of the UK. Although the heterogeneity was quite high, this was to be expected with observational studies that had a very large number of participants [31]. The heterogeneity was explored by conducting a subgroup analysis and a sensitivity analysis. The statistical analysis was methodical and thorough. Also, several outcomes of morbidity were considered. This review only included studies with the correct ethnic categorization, so that the differences amongst the Asian and MO groups would be clear. The research was limited to strictly confirmed COVID-19 cases which enhanced the validity of the SR. The search strategy was improved upon by consulting with specialist librarians at PHE and the University of Manchester. Separate adjusted and unadjusted risk estimates enhanced the understanding of the situation. The study did not include studies with overlapping participants. This review was based on UK-only populations, so the results of this review are more generalizable to the UK. Including unpublished studies in the review helped to analyze the rapidly evolving COVID-19 pandemic and improved the quality of this review.

### 4.2 Limitations

A broad ethnic classification was used in this review. The Asian group included very diverse subgroups, each of which has now been shown to have different risk profiles. This approach was used to include a wide study base, as there were very few studies that had data on sub-categories, which resulted in an incomplete assessment of the risk faced by these subgroups [5]. The search had to be limited to the UK using search terms like ‘UK’, ‘England’, and so some studies might have been missed due to this strategy. As the research was carried out during the pandemic, there was a large amount of missing data reported which hindered the analysis. This review was limited to adults and so the results could not be generalized to children. The PCR test for COVID-19 has a high false-negative rate, which led to some cases being wrongly classified as non-COVID [23]. However, this was a limitation of the UK testing strategy, rather than of this review. As this review concentrated on UK based studies, the results were less generalizable to other countries as there might be differences in hospitalization policies, testing, ICU facilities, and other factors. It has been noted that many participants’ ethnicities have been put down as ‘Other’, and this may have created erroneous results for the MO group [21]. Again, this is an error of data collection by the original study researchers, rather than of this review.

### 4.3 Policy implications

These results have urgent implications for formulating the COVID-19 response strategy including vaccination provision, protecting the BAME community which is at most risk from the worst outcomes related to COVID-19, and addressing the long-standing health inequalities.

### 4.4 Implications for research

This review needs to be upgraded to a living SR, so that the changing pandemic risks can be identified in the various ethnicities as the pandemic progresses. There is a need for more epidemiological population-based studies to assess the true risk experienced by the various ethnic groups with regards to the worst clinical outcomes.

The dataset needs to be large enough to appreciate the risk in the various subgroups.

### 4.5 Conclusion

It can be concluded that the Black, Asian, and MO groups faced the worst outcomes with regards to COVID-19 in the UK. These findings are of immense public health importance and should be used to help formulate policy concerning COVID-19 and reducing socioeconomic disparities. Racism and pre-existing disparities with regards to the wider social determinants of health are the root cause of these inequalities and need to be tackled urgently.

## Data Availability

All data presented in this review are collected from previously published and unpublished (pre-print) papers and are available from the cited references.

https://www.crd.york.ac.uk/prospero/display_record.php?ID=CRD42021248117

## Acknowledgements

Any opinions expressed in this study, only represent the views of the authors and were not necessarily related to the United Kingdom Health Security Agency. No conflict of interest exists in the submission of this manuscript. I would like to thank my family especially my sister Hina Siddiq who funded my studies, Sarah Siddiq who edited this article and my children who put up with me whilst I worked on this paper. I am also indebted to my line manager Sarah Dowle for supporting me in this endeavour.

Librarians at the University of Manchester and Public Health England (PHE) helped to formulate the search strategy. Librarians from PHE also helped to find the full text of articles.

## Notes

**Conflict of interest:** None.

**Financial Support:** This research received no specific grant from any funding agency, commercial or not-for-profit sectors.

### Competing Interest Statement

The authors have declared no competing interest.

### Funding Statement

This study did not receive any funding.

### Author Declarations

This study involves only openly available human data, which can be obtained from cited references.

