## Supplementary material S1,Supplementary table SI for "Ethnicity and outcomes in COVID-19 in the United Kingdom: a systematic review and meta-analysis"

**SUPPLEMENTARY MATERIAL S1: NOS CHECKLIST [1]**

**Longitudinal cohort studies**

A. Selection

1. Representativeness of the exposed cohort
   a. Truly representative of the average COVID-19 patients in the community (*)
   b. Somewhat representative of the average COVID-19 patients in the community, including paediatrics, hospitalised patients, etc. (*)
   c. Selected group of users (e.g., nurses, volunteers, children, healthcare workers)
   d. No description of the derivation of the cohort
2. Selection of the non-exposed cohort
   a. Drawn from the same community as the exposed cohort (*)
   b. Drawn from a different source
   c. No description of the derivation of the non-exposed cohort
3. Ascertainment of exposure
   a. Secure record (e.g., medical records) (*)
   b. Structured interview (*)
   c. Written self-report
   d. No description
4. Demonstration that outcome of interest was not present at the start of study (*)

B. Comparability

Comparability of cohorts on the basis of the design or analysis

a. Study controls for age (*)
b. Study controls for at least one comorbidity (*)

C. Outcome

1. Assessment of outcome
   a. Independent blind assessment stated in the paper, or confirmation of the outcome by reference to medical records (*)
   b. Record linkage (e.g., identified through ICD codes on database records) (*)
   c. Self-report – no reference to original medical records to confirm the outcome
   d. No description
2. Was follow-up long enough for outcomes to occur (*)

Follow-up period was adequate when it fulfilled these cut-offs:

- Mortality: 14 days [2]
- ICU admission: 9 days [2]
- Advanced respiratory support: 9 days [2]

3. Adequacy of follow up of cohorts
   a. Complete follow up – all subjects accounted for (*)
   b. Subjects lost to follow up unlikely to introduce bias – small number lost (>80% were accounted for) (*)
   c. Follow up rate <80% and no description of those lost
   d. No statement

**Supplementary Table S1: Excluded studies**

| Aldridge et al., 2020 | Wrong study type...cross-sectional study |
| --- | --- |
| Atkins et al., 2020 | Same database...UK Biobank |
| Ayoubkhani et al., 2021 | Includes suspected COVID-19 cases |
| Bannaga et al., 2020 | Full text not available |
| Boddington et al., 2021 | Risk of Infection study |
| Baumer et al., 2020 | No relevant outcomes, OR/RR/HR not given |
| Brendish et al., 2020 | Risk of Infection study |
| Brill et al., 2020 | No relevant outcomes, OR/RR/HR not given |
| Cheng et al., 2021 | Includes suspected COVID-19 cases |
| Cheng et al., 2020 | Full text not available |
| Clough et al., 2021 | Includes suspected COVID-19 cases |
| Corcillo et al., 2021 | Limited ethnic data |
| Davies et al., 2021 | No relevant outcomes, OR/RR/HR not given |
| De Lusignan, Joy M. et al., 2020 | Includes suspected COVID-19 cases |
| De Lusignan, Dorward J., et al., 2020 | Risk of Infection study |
| Dennis et al., 2021 | Includes suspected COVID-19 cases |
| Desai et al., 2020 | Full text not available |
| Drozd et al., 2021 | No relevant outcomes, OR/RR/HR not given |
| Elliott et al., 2021 | Includes suspected COVID-19 cases |
| Gates et al., 2020 | Full text not available |
| Galloway et al., 2020 | Limited ethnic data |
| Goodacre et al., 2020 | No relevant outcomes, OR/RR/HR not given |
| Ho et al., 2020 | Risk of infection |
| Hull et al., 2020 | Includes suspected COVID-19 cases |
| Joy et al., 2020 | No relevant outcomes, OR/RR/HR not given |
| Ken-Dror et al., 2020 | Limited ethnic data |
| Khalil et al., 2020 | No relevant outcomes, OR/RR/HR not given |
| Knight et al., 2020 | Includes suspected COVID-19 cases |
| Lassale et al., 2020 | Same database...UK Biobank |
| Martin et al., 2020 | Risk of Infection study |
| Miles et al., 2020 | Includes suspected COVID-19 cases |
| Milln et al., 2021 | Includes suspected COVID cases |
| Moret et al., 2021 | Full text not available |
| Nafilyan et al., 2021 | includes suspected COVID-19 cases |
| Navaratnam et al., 2021 | Includes suspected COVID-19 cases |
| Niedzwiedz et al., 2020 | Same database...UK Biobank |
| Patel et al., 2021 | includes suspected COVID-19 cases |
| Raharja et al., 2020 | Not UK based, global study |
| Raisi-Estabragh et al., 2020 | Includes suspected COVID-19 cases |
| Razieh et al., 2021 | Same database...UK Biobank |
| Richards-Belle et al., 2020 | Includes suspected COVID-19 cases |
| Sattar et al., 2020 | Includes suspected COVID-19 cases |
| Shah et al., 2020 | No relevant outcomes, OR/RR/HR not given |
| Soltan et al., 2021 | Full text not available |
| Sze et al., 2020 | Not UK based, global study |
| Tay et al., 2020 | Full text not available |
| Thompson et al., 2020 | Included Chinese in Asian group |
| Williamson et al., 2020 | Includes suspected COVID-19 cases |
| Zakeri et al., 2020 | Included Chinese in Asian group |
